# Pre-Pandemic COVID-19 in New York City: A descriptive analysis of COVID-19 illness prior to February 29, 2020

**DOI:** 10.1101/2022.04.11.22273719

**Authors:** Page Keating, Jessica Sell, Judy Chen, Joel Ackelsberg, Winfred Wu, Benjamin Tsoi, Don Weiss

**Affiliations:** New York City Department of Health and Mental Hygiene, Long Island City, Queens, New York, USA

**Author notes:** Corresponding author: Page Keating.

**Keywords:** COVID-19, COVID-19 Testing, New York City, surveillance, pandemic

## Abstract

**Background:** On January 30, 2020 the COVID-19 pandemic was declared a Public Health Emergency of International Concern (PHEIC) by the World Health Organization. Almost a month later on February 29, 2020, the first case in New York City (NYC) was diagnosed.

**Methods:** Three-hundred-sixty persons with COVID-like illness was reported to the NYC Department of Health and Mental Hygiene (DOHMH) before February 29, but 37 of these tested negative and 237 were never tested for SARS-COV-2. Records of 86 persons with confirmed COVID-19 and symptom onset prior to February 29, 2020 were reviewed by four physician-epidemiologists. Case-patients were classified as likely early onset COVID-19, or insufficient evidence to determine onset. Clinical and epidemiological factors collected by DOHMH and supplemented with emergency department records were analyzed.

**Results:** Thirty-nine likely early onset COVID-19 cases were identified. The majority had severe disease with 69% presenting to an ED visit within 2 weeks of symptom onset. The first likely COVID-19 case on record had symptom onset on January 28, 2020. Only 7 of the 39 cases (18%) had traveled internationally within 14 days of onset (none to China).

**Conclusions:** SARS-CoV-2 and COVID-19 was in NYC before being classified as a PHEIC, and eluded surveillance for another month. The delay in recognition limited mitigation effort and by the time that city and state-wide mandates were enacted 16 and 22 days later there was already community transmission.

**Key Points:** Records of 86 persons with confirmed COVID-19 and symptom onset prior to February 29, 2020 were reviewed for likelihood of early onset COVID-19. Thirty-nine likely early onset COVID-19 cases were identified, suggesting that early COVID-19 transmission in NYC went undetected.

## Background

Severe acute respiratory syndrome coronavirus 2 (SARS-CoV-2), the virus that causes COVID-19, was declared a Public Health Emergency of International Concern by the World Health Organization (WHO) on January 30, 2020 (1). The first case of COVID-19 was diagnosed in New York City (NYC) on February 29, 2020 (2, 3). In January and most of February 2020, COVID-19 diagnostic testing was only available at the U.S. Centers for Disease Control and Prevention (CDC) and was restricted to patients with severe illness, known exposure, or travel history from affected areas (4). The exponential spread of COVID-19 in March 2020 prompted the NYC Department of Health and Mental Hygiene (DOHMH) to suspect that multiple introductions and sustained transmission had occurred in January and February of 2020 (2). In this report, we review medical and investigation records of case-patients with confirmed COVID-19 who reported symptom onset prior to February 29, 2020. The objective of this analysis was to identify COVID-19 cases that might have been diagnosed before February 29, 2020, had sufficient diagnostic resources been available, to inform surveillance testing criteria in advance of the next pandemic.

## Methods

NYC residents reported to the DOHMH with confirmed COVID-19 and symptom onset between January 1 and February 29, 2020 were eligible for the study. Symptom onset date was obtained through case investigations when providers contacted DOHMH seeking laboratory testing for suspect cases of COVID-19. COVID-19 was considered confirmed when case-patients either had a positive molecular SARS-CoV-2 laboratory report received electronically through the New York State Electronic Clinical Laboratory Reporting System (5), or for deceased individuals had COVID-19 listed as a cause of death on the death certificate. Probable cases include those who tested positive for SARS-CoV-2 by antibody (one case-patient with MIS-C), or those who were symptomatic contacts of confirmed cases without laboratory confirmation of disease. Patient demographics, symptom information, comorbidities, epidemiological risk factors for COVID-19 exposure (i.e., travel history, known exposure, high-risk occupation^*^), and illness severity indicators and outcomes (i.e., hospitalization, intensive care, death) were collected by DOHMH staff. Supplemental clinical information was collected from emergency department (ED) records.

Confirmed cases of COVID-19 with reported symptom onset between January 1 and February 29, 2020 were reviewed by four COVID-19 response physician-epidemiologists. Case-patients were categorized as either likely to have had COVID-19 onset prior to February 29, 2020, or with insufficient evidence to determine early case infection status (prior to February 29, 2020). We considered initial symptoms; date of symptom onset; clinical findings on presentation to medical providers, if available; date of first positive molecular^†^ SARS-CoV-2 test, and time lapse between symptom onset and diagnostic test. If symptoms occurred 21 days or less before the positive diagnostic test, this was considered suggestive of the onset date of COVID-19. If the physician-epidemiologists agreed independently with the infection status designation, case-patients were classified accordingly. All physician-epidemiologist discrepancies were adjudicated through deliberation amongst all four physician-epidemiologists. When consensus was not reached, case-patients were assigned to the “insufficient evidence” category.

We describe case-patient demographics, epidemiological risk factors, and symptom-based classification meeting the COVID-19 Council for State and Territorial Epidemiologists (CSTE) case definition for COVID-19-like-illness (CLI)(6). Analyses were conducted using SAS software (version 9.4; SAS Institute) and R (version 3.6.3; The R Foundation).

## Results

We identified 360 individuals who were reported to the DOHMH before February 29, 2020 with CLI. Thirty-seven case-patients tested negative for SARS-CoV-2 and 237 were never tested. The remaining 86 were designated for physician-epidemiologist review; 39 (45%) were categorized as likely having had COVID-19 with onset prior to Feb 29, 2020, with the remainder (n=47) having insufficient evidence to confirm early case infection status (Figure 1). The case-patients with likely early onset COVID-19 (n=39) had a median age of 52 years (IQR 39-64). Most (56%) were male and resided in Brooklyn (33%), Manhattan (23%), Queens (21%), Bronx (18%), and Staten Island (5%). Thirty-three percent were White, 15% Black, and 33.3% Hispanic. Twenty-three percent worked in high-risk occupations and 18% reported international travel in the two weeks prior to onset, though none from China (Table 1).

**Table:** Demographics and clinical information for likely cases of COVID-19 and case-patients with insufficient evidence to determine COVID-19 status.

|  | **Likely Case of**<br>**Early Onset**<br>**COVID-19** | **Insufficient**<br>**Evidence to**<br>**Determine** |
| --- | --- | --- |
| **N (%)** | 39 (45%) | 47 (55%) |
| Died | 13 (15%) | 19 (22%) |
| **Age (median, IQR<sup>±</sup>)** | 52 (39 – 64) | 58 (45 – 63.5) |
| **Gender** |  |  |
| Male | 22 (56%) | 20 (43%) |
| Female | 16 (41%) | 27 (57%) |
| Transgender | 1 (3%) | 0 (0%) |
| **Race / Ethnicity** |  |  |
| Asian | 5 (13%) | 2 (4%) |
| Black / African American | 6 (15%) | 11 (23%) |
| Hispanic / Latino | 13 (33%) | 24 (51%) |
| White | 13 (33%) | 8 (17%) |
| Other | 0 (0%) | 1 (1%) |
| Unknown | 2 (5%) | 1 (1%) |
| **Borough** |  |  |
| Bronx | 7 (18%) | 12 (26%) |
| Brooklyn | 13 (33%) | 16 (34%) |
| Manhattan | 9 (23%) | 4 (8%) |
| Queens | 8 (21%) | 14 (30%) |
| Staten Island | 2 (5%) | 1 (2%) |
| **Met the CSTE<sup>‡</sup> definition of CLI<sup>§</sup>** | 37 (95%) | 39 (83%) |
| Fever | 34 (92%) | 29 (69%) |
| Cough | 33 (89%) | 26 (62%) |
| Shortness of breath | 21 (57%) | 24 (57%) |
| Myalgia | 17 (46%) | 16 (38%) |
| Chills | 15 (41%) | 16 (38%) |
| Loss of taste or smell | 1 (3%) | 7 (17%) |
| Congestion or runny nose | 8 (22%) | 5 (12%) |
| Diarrhea | 15 (41%) | 8 (19%) |
| Nausea or vomiting | 2 (5%) | 5 (12%) |
| **Clinical Indications** |  |  |
| Acute Respiratory Distress Syndrome | 8 (21%) | 4 (9%) |
| Pneumonia | 19 (49%) | 11 (23%) |
| **ED<sup>□</sup> Visit within 2 weeks of symptom onset** | 27 (69%) | 15 (32%) |
| Presented with CLI | 19 (49%) | 5 (11%) |
| **Other Risk Factors** |  |  |
| Travel outside of country ≤14 days of onset | 7 (18%) | 1 (2%) |
| High-risk occupation | 9 (23%) | 4 (9%) |
| **Days between Onset and Diagnosis Date** |  |  |
| Median days (IQR) | 16 (10-28) | 54 (35-86) |

<sup>±</sup> Interquartile range
<sup>≠</sup> Council of State and Territorial Epidemiologists
<sup>§</sup> COVID-19-like illness
<sup>□</sup> Emergency department

**Figure 1:**
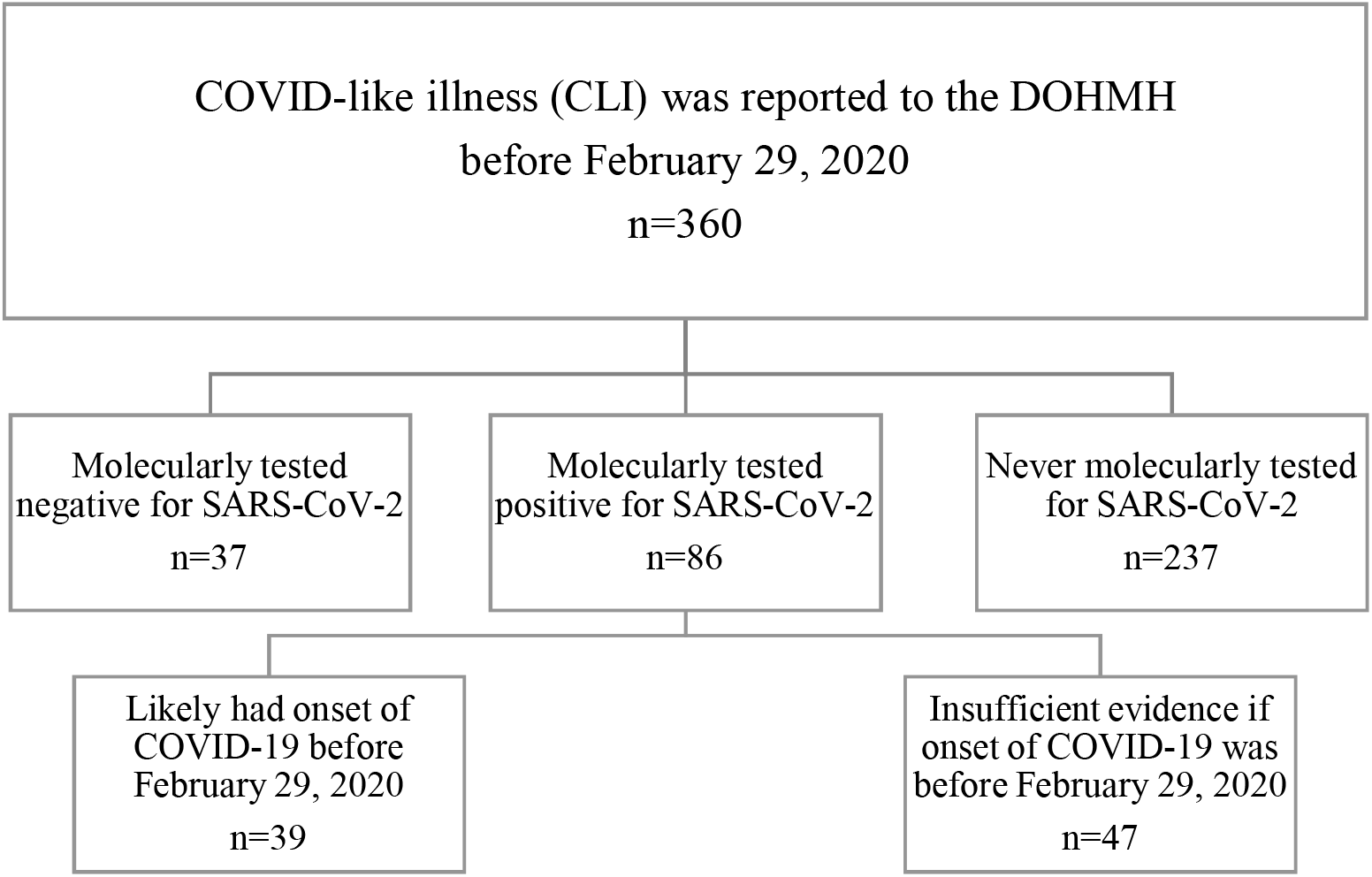
Case classification for Medical Epidemiological Review

Illness onset for the earliest likely COVID-19 case-patient was January 28, 2020, and most (69/86, 80%) who met review criteria had onset in the last 2 weeks of February (Figure 2). Common symptoms reported by case-patients were fever (92%), cough (89%), shortness of breath (57%), myalgia (46%) and chills (41%). Twenty-seven (69%) of the patients had either pneumonia (19 case-patients) or acute respiratory disease syndrome (ARDS, 8 case-patients) (Table 1). Nearly all likely early onset case-patients met the CSTE definition for CLI (95%, Table 1). Approximately half (49%) presented with CLI to an NYC ED within two weeks of symptom onset. The average number of days between symptom onset and diagnosis date was 16 days (IQR: 10-28) for likely early onset COVID-19 case-patients and 54 days (IQR: 35-86) for those with insufficient evidence to characterize COVID-19 status. The average number of days between diagnosis and ED visit was 17.5 days (IQR: 0-29) and 35 days (IQR: 0-66), respectively for likely COVID-19 cases and for those with insufficient evidence to characterize.

**Figure 2:**
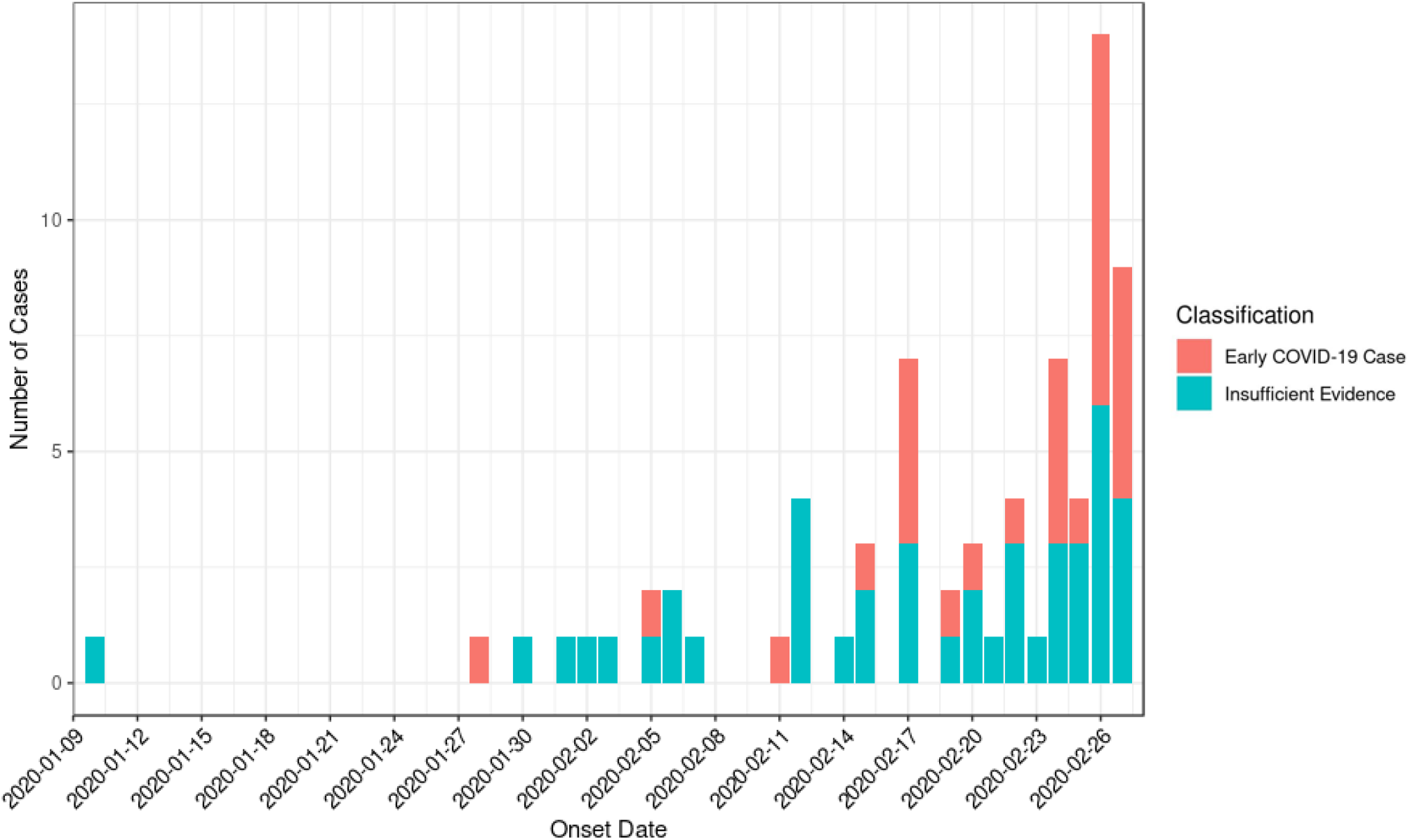
Suspected early onset cases of COVID-19 by symptom onset date and early onset case classification (likely early onset vs insufficient evidence).

Additionally, case-patients for whom there was insufficient evidence to verify their onset date more often were women, Black or Hispanic, resided in the Bronx or Queens, and had not traveled or worked in a high-risk occupation.

## Discussion

This analysis expands upon a previous DOHMH report and provides insight into the timing of NYC’s first COVID-19 wave, which caused over 200,000 cases and 18,000 deaths (2). The demographic characteristics of the likely early onset COVID-19 cases mirror those of the NYC population as does the distribution of cases across the five boroughs. A total of 360 individuals were reported to DOHMH with COVID-19-like symptoms, and the earliest case-patient with likely SARS-CoV-2 infection had onset on January 28, 2020. This person had not traveled and worked in a high-risk occupation. While our retrospective analysis identified 39 likely cases of COVID-19 prior to February 29, there undoubtedly were many more undetected COVID-19 cases in NYC. Only the most severely ill suspected COVID-19 cases, as reflected by the 69% with either pneumonia or ARDS, were tested with the newly available diagnostic test. Of the 360 individuals reported to DOHMH during the time period, 237 were not tested.

These findings illustrate that community transmission was occurring at least a month prior to recognition of the first case and indicate that the surveillance strategy was inadequate. Supporting evidence for this conclusion comes from a NYC hospital network that tested stored respiratory specimens for SARS-CoV-2 and demonstrated that the virus was likely circulating in late January 2020 or possibly earlier (7, 8).

The shortage of SARS-CoV-2 diagnostic laboratory supplies early in the pandemic hindered surveillance efforts. CDC’s criteria limited testing to persons who had evidence of lower respiratory illness and travel to Wuhan City, China (9). Since a small proportion of COVID-19 cases would have resulted in hospitalization, most would not have met criteria and been undetected (10). We, and other cities, endeavored in February 2020 to collect, and CDC agreed to test, pooled influenza negative specimens from emergency department visits in order to determine whether SARS-CoV-2 was circulating in NYC; however, we were unable to begin specimen collection until early March 2020 (11, 12).

Investments to improve federal capacity to quickly manufacture and distribute diagnostic test kits are a priority, but we contend that the approach to pre-pandemic surveillance also needs to be reconsidered. When a pandemic virus emerges, public health creates a surveillance case definition to identify suspect cases for investigation. Case definitions are often crafted to identify patients with severe disease because they are most likely to come to medical attention. Conventional public health wisdom has been to assume that initial cases will have traveled from the geographic area of origin. However, if mild, non-specific disease predominates, and the virus spreads quickly, as occurred with SARS-CoV-2, a surveillance strategy of severe illness in travelers will likely miss the early introduction. Surveillance criteria should be broadened based on available epidemiology data to detect the entire spectrum of disease of a novel virus’s entry into a population. Although the preliminary epidemiology data from China was sparse, a respiratory pathogen with person-to-person transmission would be expected to affect individuals with frequent face-to-face contact with others, especially contact with persons reporting recent travel. With the ease and speed of international travel areas of the world other than the initial outbreak zone should be included in surveillance criteria. The importation of SARS-CoV-2 into NYC has been linked to strains circulating in European countries (7).

If, or perhaps when, another novel virus emerges there again may be periods of limited testing capacity. When distributing a limited resource, such as diagnostic testing, consideration must also be given to vulnerable populations and those who historically have had inadequate access to care (13). Additionally, provisions should be made for pre-designated sentinel sites to collect specimens for pooled testing or appropriately stored for later analysis. Wastewater surveillance holds promise of early pandemic awareness if it can reliably detect and identify sequences of both known and novel pathogens (14).

Our review was limited by recall bias. Case-patients reviewed were often tested, interviewed, or went to the ED weeks after their symptom onset. Therefore, we adopted a conservative approach to classification, requiring verification of onset dates by medical or hospitalization records, to strengthen the plausibility of a single, continuous illness between symptom onset and diagnosis. All those classified as early COVID-19 cases were diagnosed with a molecular SARS-CoV-2 test. Although the blueprint for diagnostic tests for SARS-CoV-2 was published by WHO within a month of the sequence publication (15), it took another month for the CDC to release its diagnostic test (10). Even by the end of February testing availability was limited, hence our study could not capture individuals who were symptomatic and either never tested or tested negative after viral clearance.

NYC closed schools on March 16, 2020 and implemented “Pause,” the closure of all non-essential business in New York State, on March 22, 2020. Whether the knowledge that SARS-CoV-2 was circulating in NYC in late January would have prompted an earlier response is speculative, however, by the time authorities implemented population-based control measures community transmission of COVID-19 was already widespread. And while we cannot be certain when the first COVID-19 case occurred in NYC, or how many introductions propelled the first NYC wave, the delay in recognition limited the effectiveness of mitigation efforts. Our study adds to the body of evidence that pandemics can grow quickly and undetected, and that our ability to identify early introductions is paramount to the control efforts (7, 8). There is little doubt that community transmission of SARS-CoV-2 accelerated quietly and rapidly in NYC without detection before the pandemic’s arrival was announced officially on February 29, 2020.

## Data Availability

All data produced in the present work are contained in the manuscript.

## Acknowledgments

Marcelle Layton, Incident Command System Surveillance and Epidemiology Section, New York City Department of Health and Mental Hygiene

## Footnotes

* High-risk occupations include those with frequent face-to-face contact with people, such as teachers, health care workers, construction workers, security workers, entertainment industry employees, clergy, conveyance drivers, and personal care services.

† One patient diagnosed with MIS-C was diagnosed by antigen

